# Equivalent physical dose (EPD) for improved precision in voxelized dosimetry

**DOI:** 10.1101/2023.05.10.23289774

**Authors:** Elliot M. Abbott, Bleddyn Jones, Borivoj Vojnovic, Nadia Falzone, Adam Turner, Katherine A. Vallis

## Abstract

**Purpose:** Equivalent dose in 2 Gy fractions (EQD2), based on the original biological effective dose (BED) equation, is frequently used to guide treatment in the clinic. This work addresses the limitations of EQD2 in the context of voxelized dosimetry, clarifies potential sources of confusion, and provides an alternative formulation for improved precision.

**Methods and Materials:** The EQD2 formula was evaluated by a simple insertion of the EQD2 dose into the BED equation. The mathematically exact form of EQD2, referred to here as equivalent physical dose (EPD), was provided by solving the linear-quadratic model BED equation for dose using the quadratic formula. The EPD derivation was compared in terms of absolute error to the EQD2 derivation, which separates the Relative Effect term from the BED equation.

**Results:** The EQD2 expression implicitly assumed a homogenous dose, demonstrating that its use in voxelized dosimetry can mislead. As an alternative formulation, EPD was shown to adhere more closely to the first principles of radiobiological modeling. An error analysis identified absolute errors from EQD2 sometimes in excess of 10%.

**Conclusions:** Assumptions in the standard EQD2 equation are inappropriate in the context of voxelized dosimetry, where voxels within a structure, such as a target volume, may receive a dose that differs from the prescribed dose. Using EPD (or BED) instead of EQD2 would address these areas of confusion. Optimizing therapy according to biological properties in this way could provide enhanced and more reliable radiobiological input to radiotherapy treatment planning.

## Introduction

Absorbed radiation dose is a major determinant of the clinical outcome of radiation therapy for cancer. However, non-linear effects exist between dose and cell kill, for single doses and in relation to dose fractionation. Radiation oncologists and medical physicists often use “Biological Effective Dose” (BED), or its derived “Equivalent Dose in 2 Gy fractions” (EQD2) in the clinic to convert between different fractionation schemes or when adjustments are needed during a course of treatment resulting from unexpected events, or in cases where retreatment is being considered.^1^ In addition, correction factors are needed to explain phenomena such as modifications of intrinsic tissue radiosensitivity, radiation quality, DNA repair rates, and repopulation effects.^2^

Significant clinical progress resulting from the application of radiobiological principles started with the acceptance of the linear-quadratic (LQ) model.^3,4^ The LQ model defines the probability of inactivating cells as a function of radiation dose, i.e. the surviving fraction (SF) given by 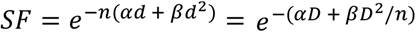 where *α* and *β* are radiosensitivity coefficients, *D* is absorbed dose given either as a single exposure or split into *n* fractions each of dose *d*. The SF is applicable to both experimental and clinical data, and is contained within the BED definition: 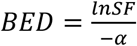. Although introducing potentially misleading units of measure, the idea of dividing the SF by −*α* allowed simpler clinical applications which required an *α*/*β* ratio as the only biological parameter. BED is expressed as^5,6*^:

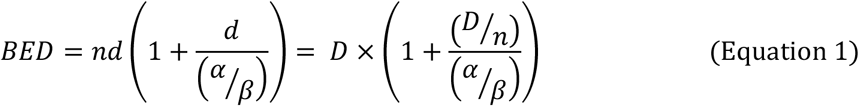

For this reason, BED (expressed in units of Gy, but followed by a suffix which denotes the operative *α*/*β* ratio) has become a valuable tool for guiding radiation therapy protocols and improving the quality of patient outcomes.

When working outside of the conventional 2 Gy fractionation schedule of external beam photon therapy, such as hypofractionated therapy at 5 Gy per fraction or more, it is sometimes helpful to convert the delivered BED into the same physical dose when given as 2 Gy fractions, as this is a fractionation schedule with which radiation oncologists are most familiar with, and for which most data concerning toxicity exists.^7^

The existing methods are well documented in the literature and rely on the concept known as equivalent dose (EQD).^3,8^ EQD is defined as the equivalent radiotherapy dose in terms of biological effects, where dose per fraction has been incorporated. EQD is commonly derived by dividing BED models by a “relative effect” (RE) term, where

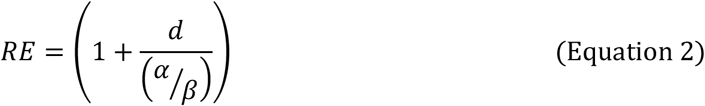

as contained in the BED equation (see Appendix A). This derivation yields the following general formula:

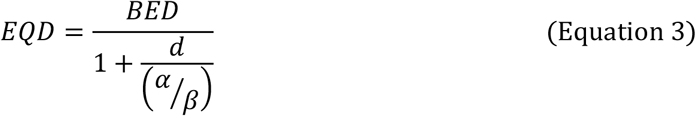

which is recommended by the Report Committee 25 of the International Commission on Radiation Units & Measurements (ICRU).^8^

In the radiation oncology community, EQD is conventionally simplified in terms of 2 Gy fractions (EQD2) by substituting *d* = 2 for 2 Gy fractions into the general EQD equation such that:^7^

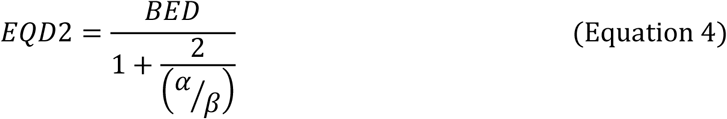

Note that the EQD2 formula may also appear as:

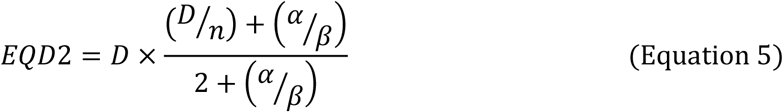

which is the same quantity reduced with the numerator and denominator multiplied by *α*/*β*. EQD2 is the most widely used tool to account for variation in treatment schedules and is suitable for the comparison of dose prescriptions, provided that there is little or no dose inhomogeneity. Where dose inhomogeneity exists, the expression of an equivalent dose in 2 Gy fractions may not be correct for a voxel which does not contain the intended dose *D*.

We demonstrate here that there are limitations to the frequently applied expressions of EQD2 in their most widely utilized forms, especially for heterogeneous doses, which can lead to voxelized dose evaluation inaccuracies in excess of 10%. Two propositions are presented. They lead to the conclusion that both the EQD2 and general EQD expressions have limitations that should be considered when designing or comparing radiation therapy protocols in voxelised forms.

- **Proposition 1:** The EQD2 expression assumes that the exact prescription dose is homogeneously delivered to all tissues, which is unrealistic particularly in the case of organs-at-risk (OARs), where dose is expected to vary from moderately high close to the tumour target to low or zero elsewhere.
- **Proposition 2:** EQD is derived without a mathematically exact definition when dose is variable, as in the case of voxelised plans that inherently contain deviations from the prescription dose. This introduces systematic errors in terms of isoeffect that increase as variation from the prescribed dose increases.

To address these propositions, this article provides an alternative isoeffective dose formulation, referred to as equivalent physical dose (EPD), which is defined from this point onwards as the inverse of a biological effective dose that returns an isoeffective physical dose, such that:

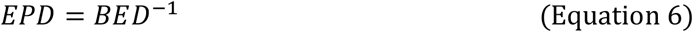

where *BED*^−1^ represents the mathematical inverse function which reverses the effects of BED calculation rather than its reciprocal or multiplicative inverse (1 ÷ *BED*). The purpose of the inversion is to derive a complementary expression to enable direct conversion from biological effect units into physical dose units by undoing the operations that evaluate biological effectiveness from the cell survival curve as a function of physical absorbed dose and other relevant treatment parameters. EPD is derived below (see Appendix B) for the LQ model and used in worked examples with an assessment of the quantitative impact of existing systematic errors.

## Methods

### Proposition 1: Proof that EQD2 does not apply to heterogeneous dose

In external beam radiotherapy (EBRT), spatially fractionated techniques like LATTICE and GRID therapies intentionally implement heterogeneous dose distributions to take advantage of hypothesized bystander/abscopal effects and immune response activation, to overcome absorbed dose constraints in the target. In conventional EBRT, depending on the method of treatment delivery, e.g., 3D conformal, static field intensity modulated radiotherapy (IMRT), or volumetric modulated arc therapy (VMAT), some voxels within a clinical target volume (CTV) or planning target volume (PTV) do not receive the prescribed target dose.^9^ Hypofractionated EBRT used for stereotactic radiosurgery (SRS) and stereotactic radiation therapy (SRT) also features highly inhomogeneous doses to target volumes in order to maximize dose gradient index in normal tissue.^10,11^

Despite the known heterogeneity of dose distributions, the standard equation 4 formula for EQD2 assumes homogeneity, which does not reflect clinical reality.^12^ EQD2 using the standard formula:

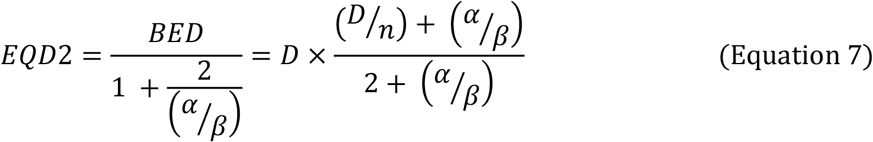

should not be used for areas of heterogeneous dose absorption. The fixed *D* in the target and the “2” in the denominator assume that the irradiated volume receives a uniform “prescription dose”. This introduces errors in real-world voxelized situations. In reality each voxel receives a dose, which often varies from the prescribed dose, and carries localized biological implications for response and toxicity. Delivering a perfectly homogeneous 2 Gy dose is practically not possible.

More importantly, fixing a dose of 2 Gy per fraction neglects the fact that the full prescription dose is not delivered to entire normal organs. An EQD2 of say *x* Gy, is only valid if *x* can be divided by 2 to form an integer. Generally, absorbed dose in normal tissues is aimed to be much lower than the prescribed dose per fraction of 2 Gy, although some normal tissues (those within the CTV and PTV) do receive the full prescribed dose and in some instances higher doses (in so-called “hot spots”) and so the risk of normal tissue complication is not adequately considered when 2 Gy is fixed in the denominator. This can be corrected by input of the correct dose for each voxel evaluated. Furthermore, the primary limitation to the maximum dose that can be delivered to tumor is the OAR dose: in other words, it would be possible to give much higher doses to the tumor if not for the risk of normal tissue toxicity. It follows that the accuracy of equivalent dose to normal tissue is critically important, and yet the standard EQD2 model implicitly does not consider this parameter. EQD2 should only be applied to specified sub-regions where dose is reasonably uniform.

### Proposition 2: Proof that EQD can be misleading

The classical derivation^5^ of EQD is given in Appendix A. However, this derivation separates the dose and relative effect (*RE*) terms, which diminishes the functionality of the LQ model. This is because *RE* contains *D*, and so separation of the two terms results in incompletely solving the equation for D. Separating D from *RE* effectively asserts a new equality (*EQD* = *D*) substituted only in the non-RE term, whereas it should simply be solving the complete equation directly for *D*. To clarify this explicitly by contrasting equations A3-A5 with B3-B5 (see Appendix A and Appendix B), one must simultaneously substitute EQD in the RE term to yield:

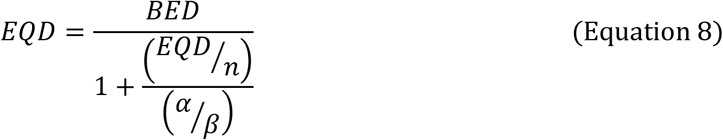

as per equation A8 of Appendix A. This paradoxically requires that EQD must be a function of itself to preserve the original behavior of the BED equation, where the referenced dose is not fixed. Critically, separating *D* from RE means that the equation 4 expression for EQD2 is not invertible.

The derivation of EPD, in physical dose units of Gy and which follows the inverted formulation (*EPD* = *BED*^−1^), is given in Appendix B. Note that this formulation is not new and has been presented previously by Fowler (see Eq 11 in reference 3)^3^. Alongside EQD, Fowler presented EPD to be required when “the number of fractions is altered instead of the dose-per-fraction”. Fowler makes no claims here about heterogeneous dose or normal tissue doses, other than that “it is useful to work out several results with different values of α/β”.

### Error analysis

The discrepancy introduced through use of the standard EQD approach may generate results that are misleading. Here, this is quantitatively defined as:

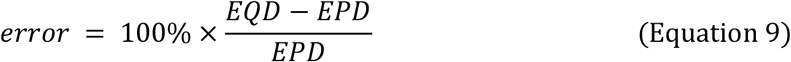

## Results

Two treatment regimens, 60 Gy in 30 fractions of 2 Gy each and 48 Gy in 16 fractions of 3 Gy each, that have the same BED if the α/β is 2 Gy (as in the central nervous system), were used in sample calculations. Table 1 shows a comparison of calculations using the equivalent dose expressions (EQD2, revised EQD2, and EPD) for these regimens.

**Table 1:** Sample calculations comparing the (1) EQD2, (2) revised EQD2, and (3) EPD calculation methods. Treatments I and II are biologically equivalent but have different total prescribed doses and number of fractions. The combinations of isoeffective dose formulae and treatment regimens here compare a prescription voxel (achieving the desired prescribed dose of 60 Gy) with a non-conforming voxel (achieving 75% of the prescribed dose). EQD2 is accurate only for a voxel receiving the prescribed dose because of the underlying assumptions of its formulation. BED for the non-conforming voxel is calculated separately from the treatment prescription BED because that voxel is absorbing a dose of radiation that differs from the prescribed dose.

| Equations in Appendix C<br>Assume $\alpha/\beta = 2 \text{ Gy}$ | **Treatment I (reference)**<br>(30 fractions at 2 Gy/fraction)<br>Prescription dose of $D_{I,Rx} = 60 \text{ Gy}$<br>$n_I = 30$ | **Treatment II**<br>(16 fractions at 3 Gy/fraction)<br>Prescription dose of $D_{II,Rx} = 48 \text{ Gy}$<br>$n_{II} = 16$ |
| --- | --- | --- |
| **Prescription voxel**<br>$D_{Rx}$ | $D_{I,Rx} = 60 \text{ Gy}$ $D_{I,Rx}/n_I = 2 \text{ Gy/fraction}$ $BED_{I,Rx} = 60 \text{ Gy} \left( 1 + \frac{60 \text{ Gy}/30}{2 \text{ Gy}} \right) = 120 \text{ Gy}$<br>[A]<br>$EQD2_{I,Rx} = \frac{120 \text{ Gy}}{1 + \frac{2 \text{ Gy}}{2 \text{ Gy}}} = 60 \text{ Gy} = D_{I,Rx}$ $\text{revised } EQD2_{I,Rx} = \frac{120 \text{ Gy}}{1 + \frac{(2 \text{ Gy} \times 30)/30}{2 \text{ Gy}}} = 60 \text{ Gy} = D_{I,Rx}$ $EPD_{I,Rx} = 30 \frac{2 \text{ Gy}}{2} \left( \sqrt{1 + \frac{4(120 \text{ Gy})}{30(2 \text{ Gy})}} - 1 \right) = 60 \text{ Gy}$ $= D_{I,Rx}$ | $D_{II,Rx} = 48 \text{ Gy}$ $D_{II,Rx}/n_{II} = 3 \text{ Gy/fraction}$ $BED_{II,Rx} = 48 \text{ Gy} \left( 1 + \frac{48 \text{ Gy}/16}{2 \text{ Gy}} \right) = 120 \text{ Gy} = BED_{I,Rx}$<br>[B]<br>$EQD2_{II,Rx} = \frac{120 \text{ Gy}}{1 + \frac{2 \text{ Gy}}{2 \text{ Gy}}} = 60 \text{ Gy} = D_{I,Rx}$ $\text{revised } EQD2_{II,Rx} = \frac{120 \text{ Gy}}{1 + \frac{(3 \text{ Gy} \times 16)/30}{2 \text{ Gy}}} = 66.67 \text{ Gy}$ $= 1.11 \times D_{I,Rx}$ $EPD_{II,Rx} = 30 \frac{2 \text{ Gy}}{2} \left( \sqrt{1 + \frac{4(120 \text{ Gy})}{30(2 \text{ Gy})}} - 1 \right) = 60 \text{ Gy}$ $= D_{I,Rx}$ |
| **Non-conforming voxel**<br>$D_{nc} = 75\% \times D_{Rx}$ | $D_{I,nc} = 75\% \times D_{I,Rx} = 0.75 \times 60 \text{ Gy} = 45 \text{ Gy}$ $D_{I,nc}/n_I = 1.5 \text{ Gy/fraction}$ $BED_{I,nc} = 45 \text{ Gy} \left( 1 + \frac{45 \text{ Gy}/30}{2 \text{ Gy}} \right) = 78.75 \text{ Gy}$ $= 0.656 \times BED_{I,Rx}$<br>[C]<br>$EQD2_{I,nc} = \frac{78.75 \text{ Gy}}{1 + \frac{2 \text{ Gy}}{2 \text{ Gy}}} = 39.375 \text{ Gy} = 0.875 \times D_{I,nc}$ $= 0.875 \times EPD_{I,nc}$ $\text{revised } EQD2_{I,nc} = \frac{78.75 \text{ Gy}}{1 + \frac{(1.5 \text{ Gy} \times 30)/30}{2 \text{ Gy}}} = 45 \text{ Gy} = D_{I,nc}$ $= EPD_{I,nc}$ $EPD_{I,nc} = 30 \frac{2 \text{ Gy}}{2} \left( \sqrt{1 + \frac{4(78.75 \text{ Gy})}{30(2 \text{ Gy})}} - 1 \right) = 45 \text{ Gy} = D_{I,nc}$ | $D_{II,nc} = 75\% \times D_{II,Rx} = 0.75 \times 48 \text{ Gy} = 36 \text{ Gy}$ $D_{II,nc}/n_{II} = 2.25 \text{ Gy/fraction}$ $BED_{II,nc} = 36 \text{ Gy} \left( 1 + \frac{36 \text{ Gy}/16}{2 \text{ Gy}} \right) = 76.5 \text{ Gy}$ $= 0.638 \times BED_{I,Rx}$<br>[D]<br>$EQD2_{II,nc} = \frac{76.5 \text{ Gy}}{1 + \frac{2 \text{ Gy}}{2 \text{ Gy}}} = 38.25 \text{ Gy} = 0.867 \times EPD_{II,nc}$ $\text{revised } EQD2_{II,nc} = \frac{76.5 \text{ Gy}}{1 + \frac{(2.25 \text{ Gy} \times 16)/30}{2 \text{ Gy}}} \approx 47.81 \text{ Gy}$ $= 1.084 \times EPD_{II,nc}$ $EPD_{II,nc} = 30 \frac{2 \text{ Gy}}{2} \left( \sqrt{1 + \frac{4(76.5 \text{ Gy})}{30(2 \text{ Gy})}} - 1 \right) \approx 44.10 \text{ Gy}$ |

In this example, the dose delivered to a hypothetical non-conforming voxel in the target volume is 75% of the prescribed dose (*Rx*) and so this voxel receives 1.5 Gy/fraction rather than the intended 2 Gy/fraction. This magnitude of non-conformity to points within the target volume is a common occurrence even with modern treatment modalities; however, the number of scheduled fractions is a factor that may be controlled exactly. Depending on the extent of the non-uniformity of the absorbed dose throughout the target, it is possible that the number of “non-conforming voxels” will exceed the number of “conforming voxels”.

In Table 1, Box A, the prescription voxel is exactly correct for all formulations of equivalent dose because they equal *D*_*Rx*_. The fact that all of these formulations yield the same correct result masks their differences for an alternative fractionation scheme or the case of non-conforming voxels, which underscores the confusion about the generalizability of BED modeling. In Box B, the prescription voxel is exactly correct for the EQD2 formulation but inexact for the revised EQD2 formulation, with a difference of +11%. In Box C, the non-conforming voxel is inexact for the conventional EQD2 formulation, differing by -12.5%, but is exactly correct for the revised EQD2 formulation. In Boxes A-C, EPD matches the expected isoeffective physical dose, either *D*_*nc*_ or 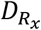. Finally, in Box D, EPD does not equate to *D*_*nc*_ nor 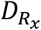 because it is simultaneously considering non-conforming voxel and change in fractionation regimen such that the BED values differ. Taking the EPD value as ground truth, the non-conforming voxel is inexact for the conventional EQD2, differing by -13.3%, and is also inexact for the revised EQD2 with a difference of +8.4%.

Because Treatment I is given in 2 Gy fractions, the method can be directly validated by comparing the calculated equivalent dose of the voxel with its original physical dose. The equivalent dose for Treatment I (2 Gy/fraction) using EQD is exact for the voxel receiving the prescribed dose. However, for a dose not conforming to the exact prescription dose, systematic errors of the EQD method of ≈±10% are demonstrated.

In Figures 1 and 2, the equivalent dose calculation errors, shown in Table 1, are presented graphically. Figure 1 illustrates a delivered dose (x-axis) and isoeffective dose (y-axis) calculation deviating from the prescription dose for regimens that (A) have the same dose per fraction as Treatment I and that (B) have a different dose per fraction, as for Treatment II. Figure 2 plots the errors shown in Figure 1 as a function of dose to demonstrate the limitations of the EQD2 model. Here, EQD2 error increases proportionately as dose deviates from the prescription dose. Conversely, the revised EQD2 formulation scales with EPD so the errors are lower for large deviations from the prescription dose.

**Figure 1:**
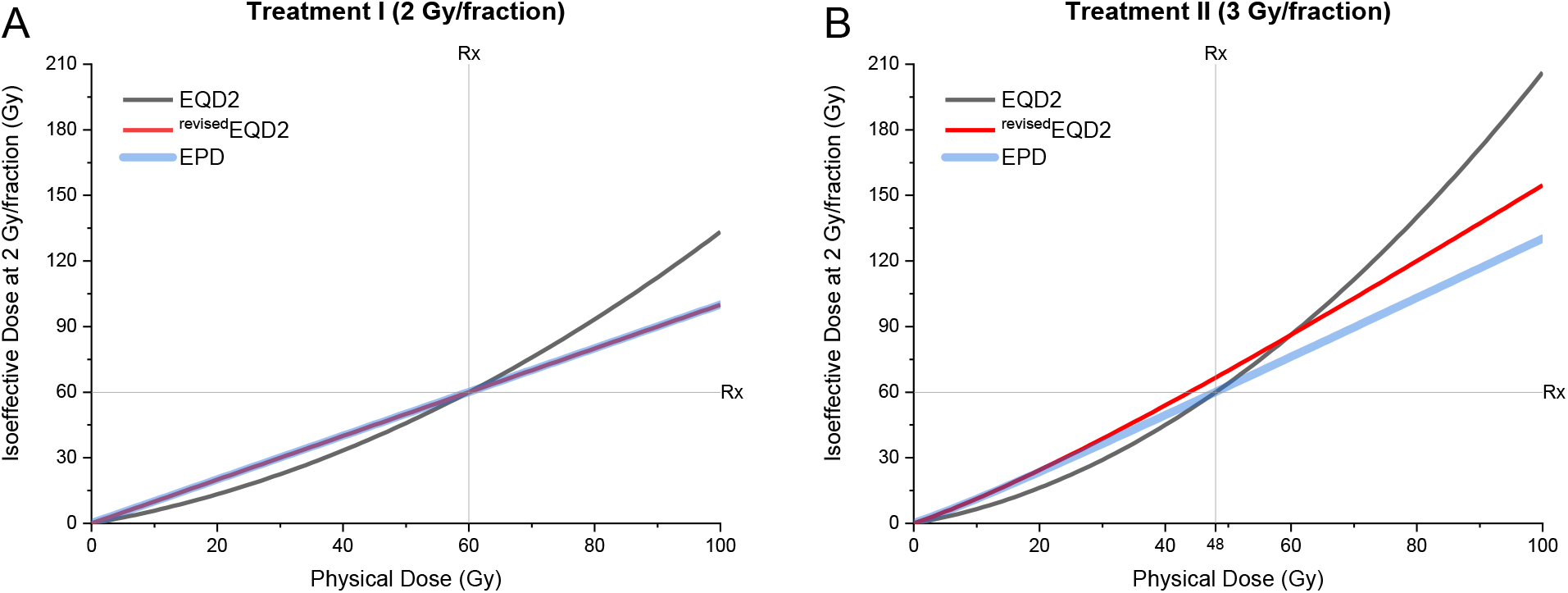
EQD2 calculations performed for two different biologically equivalent treatment schedules with *BED* = 120 *Gy* at the prescribed dose. (A) Treatment I delivered 60 Gy in 30 fractions of 2 Gy each. All equivalent dose equations compute the same EQD2 value of 60 Gy as the prescribed dose, which is as expected because treatment was delivered in 2 Gy fractions. The conventional EQD2 equation deviates from identity and therefore incorporates error for non-conformal voxels in the treatment volume. The revised EQD2 and EPD equations have a slope of unity, since the regimen is set at 2 Gy/fraction. (B) Treatment II delivered 48 Gy in 16 fractions of 3 Gy each. The revised EQD2 equation has a different slope than the EPD equation and the conventional EQD2 equation returns the expected isoeffective dose only at the prescribed dose

**Figure 2:**
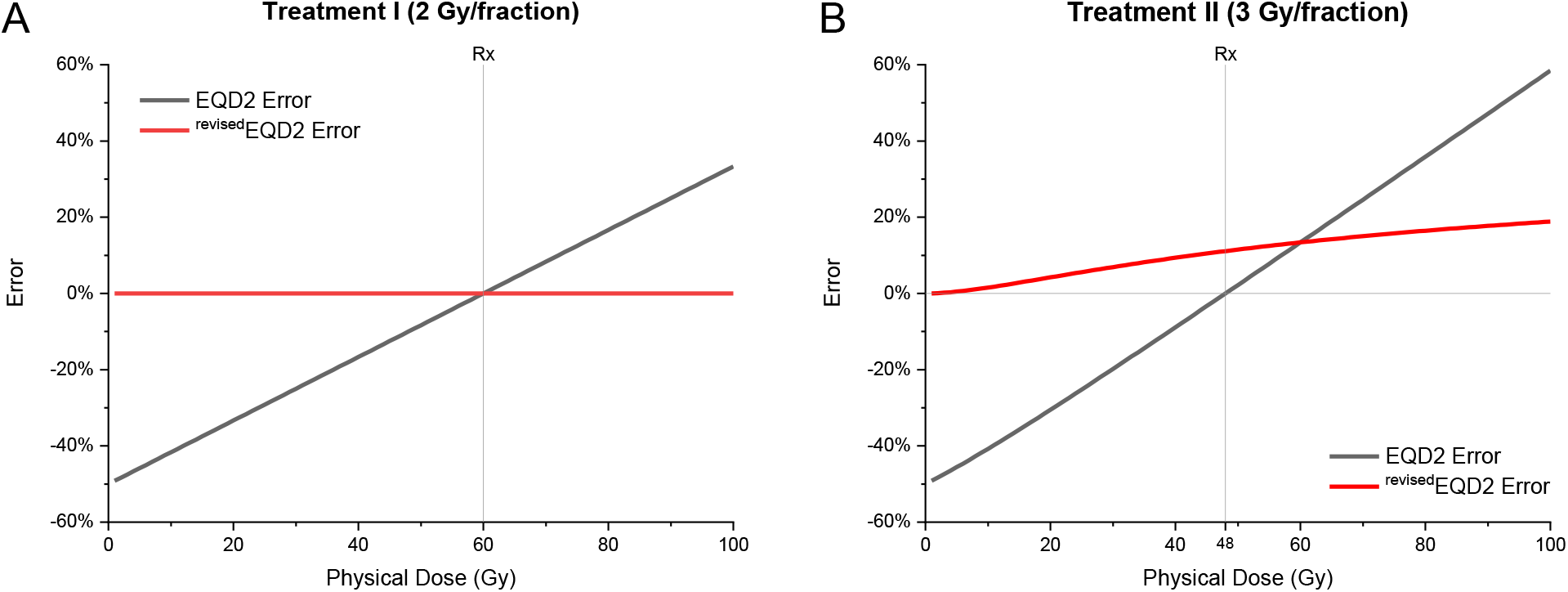
Error introduced from the formulations of EQD2 relative to the EPD equation. (A) The conventional EQD2 formulation results in 5% error for every 6 Gy of deviation from the prescribed dose when converting Treatment I schedule of 2 Gy/fraction to EQD2. (B) The EQD2 formulation has 0% error at the prescribed dose and an additional 1.1% error for every Gy deviation from the prescribed dose when converting the Treatment II schedule of 3 Gy/fraction to EQD2.

As shown in Figure 2, it is possible that EQD modeling introduces much higher errors than the approximately ±10% example in Table 1 as dose varies from the prescription dose. Furthermore, to account for uncertainties in the LQ model parameter estimates due to treatment site, tumor histology, and the applied LQ model, it is recommended to explore a range of parameter (e.g., α/β) values.^13^ Figure 3, demonstrates errors in excess of 50% depending on the set of radiobiological parameters and doses involved. The error introduced per Gy of deviation from the prescribed dose is greater for smaller values of α/β. Since normal tissues typically have smaller α/β ratio than do tumors, the EQD2 errors are most likely to apply to normal tissues and to late effects.

**Figure 3:**
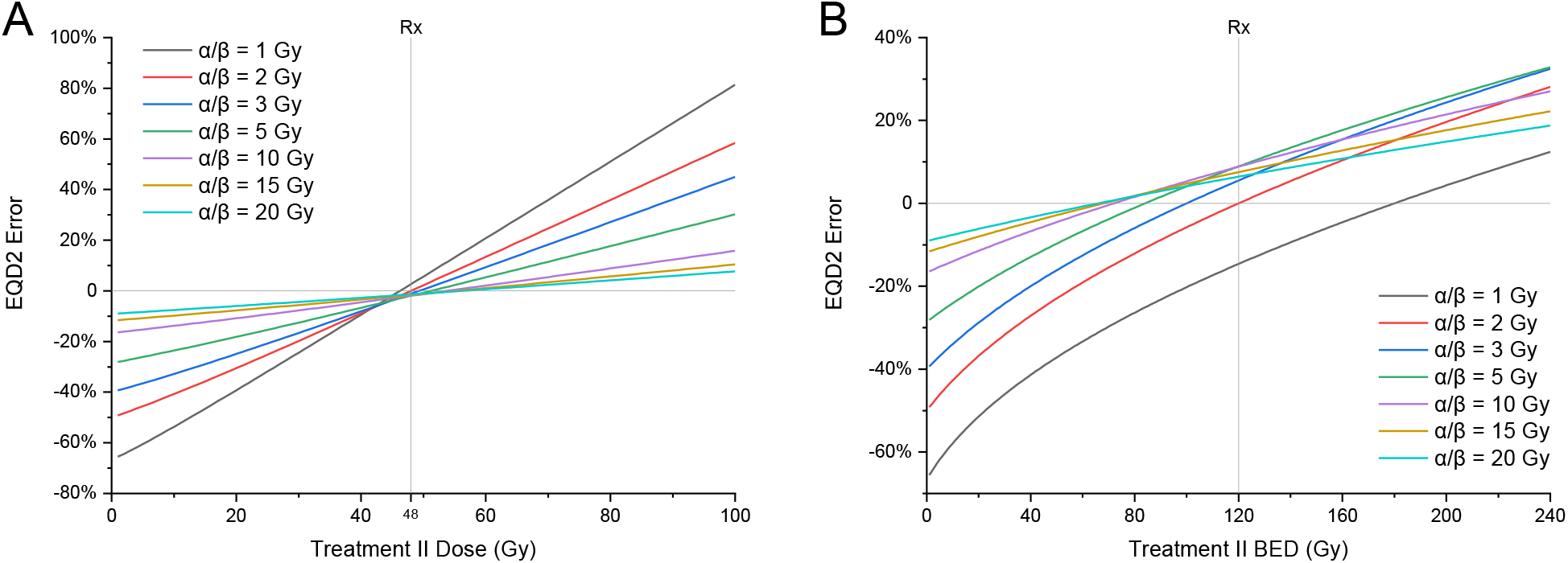
Errors in EQD2 of Treatment II but with variable α/β ratios. (A) The slope of the error in EQD2 versus Treatment II physical dose increases as the α/β ratio decreases from 20 to 1 Gy. (B) the error in EQD2 versus Treatment II BED varies with the α/β ratio. Note that, because Treatment I (2 Gy/fraction) is not isoeffective (i.e., *BED* ≠ 120 *Gy*) for different α/β ratios, prescribed dose shows 0% error only for *α*/*β* = 2 *Gy*, which was assumed throughout this work. However, if instead fixing *BED* = 120 *Gy* and allowing the number of fractions to vary with α/β ratio, the relative number of fractions increases for increasing α/β ratio such that, for*α*/*β* = 3 *Gy*, Treatment II (*d* = 3) would need to deliver 10 fractions while Treatment I (*d* = 2) delivered 36 fractions to achieve isoeffective schedules with *BED* = 120 *Gy*, so a difference of 26 fractions instead of 14 just by changing α/β from 2 Gy to 3 Gy.

## Discussion

In the current era of voxelized dosimetry, prescribed dose-only based BED calculations that assume a uniform dose may generate misleading results. Ideally, radiobiological effects resulting from variation of dose in the tumor and normal tissue volumes should be considered. A framework to comprehensively characterize the biological effects of absorbed doses in a voxelized manner is presented here. It may also be suggested that the definition of EQD2 should be made more precise, for example “EQD2 is the equivalent dose given in 2 Gy fractions to achieve a specified biological effect if (and only if) there is dose homogeneity. Additional measures must be taken if this is not so.”

In any radiotherapy plan there are likely to be non-conformal voxels that receive more or less dose than the intended (prescribed) dose to a specific tumour or normal organ. In some cases, it is estimated from the results in Figure 2, these deviations could introduce errors >50% in EQD2, with further fluctuations due to biological parameter values, such as those for *α*/*β*. Significant deviations may be most common in treatment techniques with steep dose gradients such as stereotactic body radiation therapy (SBRT), where dose decreases nearly isotropically in and around the target,^14^ or high dose rate (HDR) brachytherapy, where absorbed dose in some voxels near the source can exceed the prescription dose by 200%.^15,16^ Overall, such as when taking the mean dose across a structure, some of these errors may cancel out and this may account for the acceptance of the existing models to date, particularly in parallel organs, where the volume effect plays a greater role.^17^ However, in serial organs such as the spinal cord, the accuracy of dosimetry at the voxel level is critical. Furthermore, because tumor control depends on eradicating 100% of tumor cells, leaving even a single voxel under-treated could adversely affect the probability of cure. Paradis *et al*. ^18^ alluded to this for re-irradiation of patients “Evaluation of biological mean dose involves the conversion of each voxel within the structure to biological dose before calculating the mean. This is problematic in that doses very different from 2 Gy per fraction may skew the mean, making the result difficult to interpret.” ^18^

The findings for Proposition 1 alone, addressed by the revised EQD2 formulation, do not fully address the shortcomings as the EPD formulation does. However, the revised EQD2 formulation is valuable in demonstrating the inaccuracy of assuming a fixed denominator of (2 + (*α*/*β*)). There is also the question of the choice of parameters as set values, e.g. 3 Gy for normal tissue and 10 Gy for tumor tissue have been assumed in most cases. In addition, the magnitude of the error introduced depends on model parameterization. Normal tissues with smaller *α*/*β* ratios will be affected more by errors introduced when using the EQD method.

Nevertheless, existing BED models and variations thereof are adequate, as long as they are mechanistically understood and have a clear evidence base (with transparent uncertainty and traceability). A published letter by Fowler^19^ in 2013 recommended against the use of EQD for regimens other than 2 Gy per fraction because of the extensive experience of and familiarity with outcomes with this fractionation scheme. The existing EQD2 might be a reasonable approximation when the assumptions addressed in this article are considered. This has been noted by Withers *et al*. where uncertainty in the specific modeling parameters for each tissue carries substantial uncertainties.^20^ For this reason, caution was advised when employing the EQD model for doses outside of the limits where radiation sensitivity has been adequately characterized. The introduction of treatment dose variations by a dose-modifying parameter *g*, was suggested^21^, based on earlier work^22^, such that *g* = 0.9 and 0.8 where there are 10 and 20% reductions from the prescribed dose in a normal tissue respectively, such that hypofractionation effects could be better assessed. This led to the concept of “treble trouble” caused by hypofractionation if not properly compensated for, beyond the usual “double trouble” when using 2 Gy fractions as noted by Withers.

Based on these results, EQD2 may not be the best approach to use and may have caused confusion in the field. To calculate iso-effect or equivalent physical dose, the BED model equation can simply be inverted, as per the EPD equation, and therefore find the expression for physical dose as a function of BED or SF. Interestingly, this inversion approach, assuming that an inverse solution exists, should also be suitable for other bioeffect models beyond the conventional LQ model, provided these directly relate physical dose to biological effect. Such models include modifications of the LQ model with additional terms that account for repopulation,^23^ relative biological effectiveness (*RBE*_*max*_), and Lea-Catcheside dose protraction factor (G).^24^ When these modifying terms of the LQ model do are not dependent on physical dose, deriving EPD is readily achievable with the inversion approach (see Supplemental Materials).

Furthermore, even when considering only the *α*/*β* ratio, these results illustrate that great care must be taken when using the EQD2 model. The Figure 3 findings, that errors introduced by EQD2 are more significant as *α*/*β* becomes smaller, mean that EQD2 calculations for late effects in normal tissues (where *α*/*β* is often smaller) is subject to relatively greater errors than for tumor tissues. Consequently, composite dose estimations using EQD2, particularly in voxelized plans, may be subject to greater quantitative uncertainties. Given the variability of clinical BED modeling parameters in literature^13^, reducing sources of systematic error in the modeling could help to further refine parameter characterization.

Several authors have emphasized the pitfalls of existing radiobiology modeling approaches and added complexity due to additional radiobiology considerations. Lee *et al*. stated^25^ “the first trouble comes from the difference between physical dose prescribed and actual dose received at any point (heterogeneity of total dose), and the second results from the variation of biological effects with different dose per fraction (heterogeneity of dose per fraction). Conscientious clinicians will often estimate such biological effects semi-quantitatively using dose-volume histograms and decide upon a particular treatment plan accordingly. Yet, one wonders if a biologically oriented treatment plan could be more precise, thus representing a useful clinical tool.” The proofs of Propositions 1 and 2 address these “troubles” directly and, with the added precision in the context of voxelised dosimetry, and suggest at the possibility of biologically oriented treatment planning.

As stated by the ICRU Report Committee on Bioeffect Modeling and Biologically Equivalent Dose Concepts in Radiation Therapy^8^ “We recognize the fact that two main schools of bioeffect modeling (EQD2 vs. BED) have developed over time. They use mathematically equivalent formalisms and the difference between them should not be exaggerated. However, differences in concepts and terminologies that may appear similar to the non-expert can cause confusion. We are therefore strongly supportive of recent calls for a consensus on how bioeffect calculations and their results should be reported. It is in everybody’s interest to agree on a consistent and standardized terminology as well as recommendations on quantities and units with which authors and users of bioeffect models should strive to comply.” The mathematical critique provided by the EPD explanation could potentially offer clarity required to guide consolidation of the EQD2 and BED schools of bioeffect modeling.

Current methods of calculating EQD2 assume uniform prescription dose-based calculations and have been successful in their ease of implementation. They are, however, flawed by the assumptions that prevent their generalized application in radiotherapy. It is advised that EQD2 not be used uncritically for voxelized dosimetry unless the underlying assumptions are fully understood. Otherwise, the equivalent dose expressions illustrated here can be used with a mathematical justification. In the era of voxelized dosimetry, the approach presented here empowers radiation oncology clinics to perform voxelized biological effective dose modeling to correctly evaluate biological effective dose accumulation for common clinical contexts. This includes re-irradiation and combination therapies like EBRT boosted with HDR brachytherapy.

## Data Availability

All data produced in the present work are contained in the manuscript

## Appendix A: Derivation of EQD

### Derivation

For two treatments, *x* and *y*, the equivalent dose of a treatment in *d*_*x*_ Gy per fraction in terms of an isoeffective treatment in *d*_*y*_ Gy per fraction would be related by the following relationship:

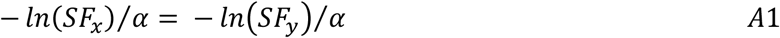

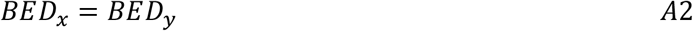

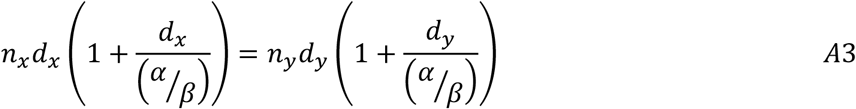

The labelling of EQD is then based on the idea that the relative effect term in *BED* = *D* × *RE* can be used to divide both sides by the same *RE*. However, this is not the intention of what the original SF equations being set equal to each other was asserting, because a new relationship *EQD*_*x*_ = *n*_*x*_*d*_*x*_ is introduced, making an assumption that the *RE*_*x*_ is not dependent on *d*_*x*_, such that both instances of *d*_*x*_ may be separated, despite the fact that both *d*_*x*_ terms are co-dependent and must be solved for simultaneously.

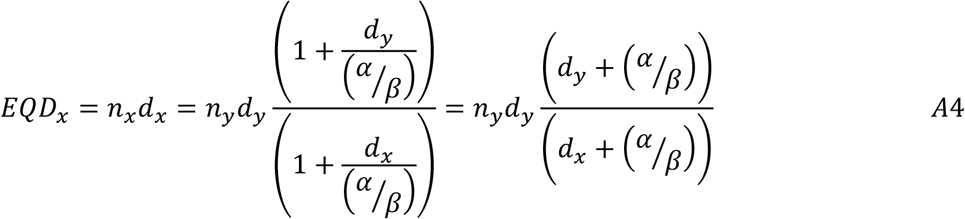

Rewriting Equation A4 with simplified notation by replacing *n*_*y*_*d*_*y*_ with *nd* yields

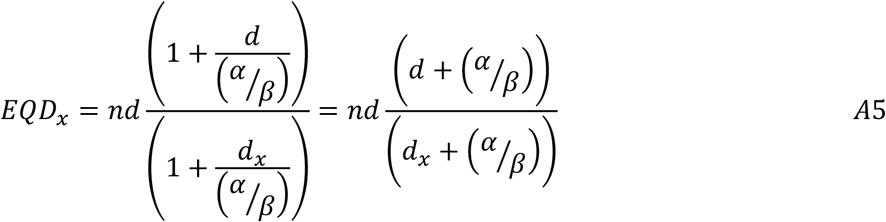

Rewriting Equation A5 with an alternative simplified notation by replacing *d* with *D*/*n* and *d*_*x*_ with *x* yields

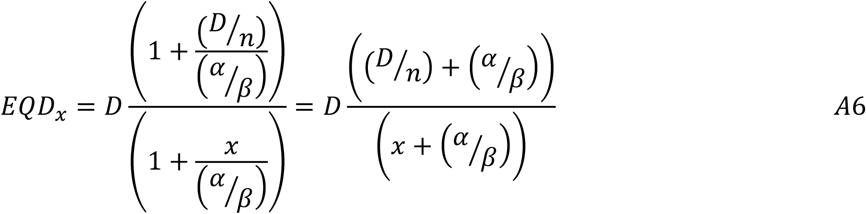

To compare *d* Gy per fraction to *d*_*x*_ = 2 *Gy* per fraction, Equation A5 reduces to the familiar EQD2 expression:

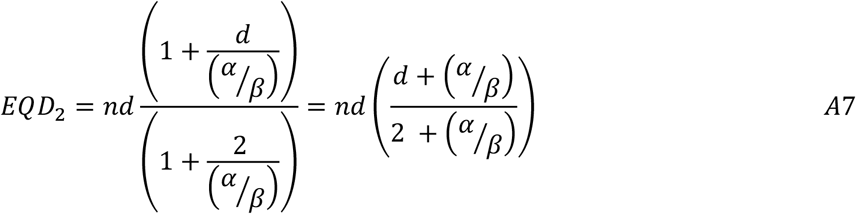

### Invalidation of equivalent biological effect

The BED of an EQD for the reference therapy should match the BED of the original therapy, i.e. be invertible. To test this, EQD can be directly inserted into the BED expression.

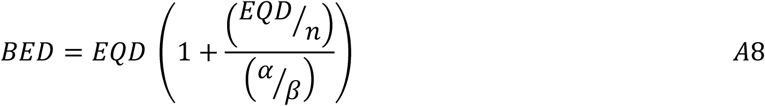

Substituting the expression for EQD from Equation A6 results in an expression which cannot be reduced.^†^

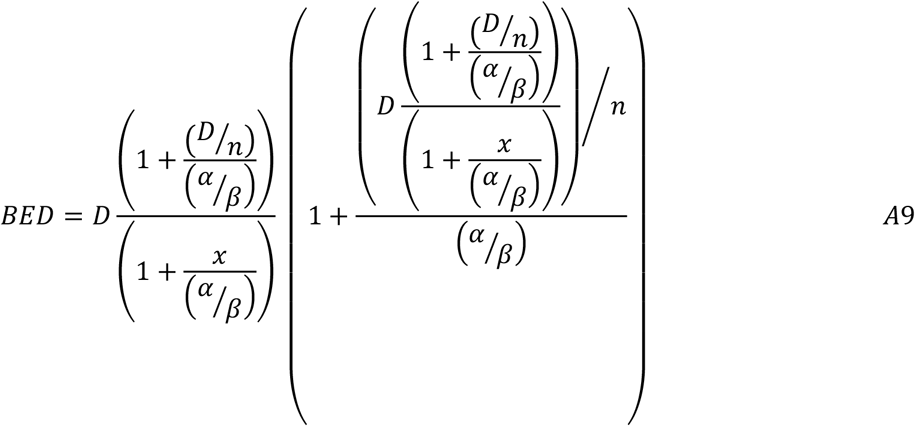

Therefore, attempting to calculate the BED of an EQD (i.e. invert the EQD operation) does not result in the same BED value from the original therapy. Rearranging the terms which appear to cancel out might seem like a possible counter proof. However, this just reproduces the “relative effect” approach described above by touching the left-hand side and treating both uses of *d* as independent when they are actually co-dependent.

## Appendix B: Derivation and validation of the EPD equation

### Derivation

The equivalent (or iso-effective) dose of treatment y in *d*_*y*_ Gy per fraction in terms of a treatment x in *d*_*x*_ Gy per fraction would be related by the following equation:

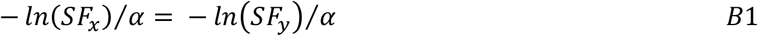

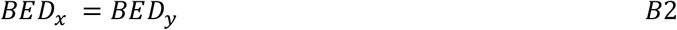

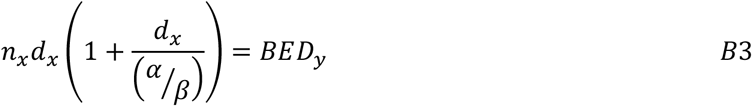

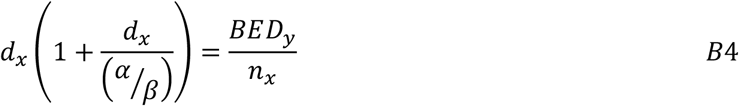

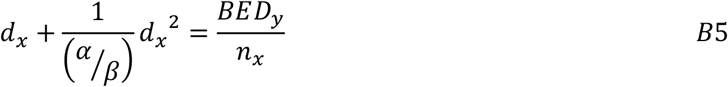

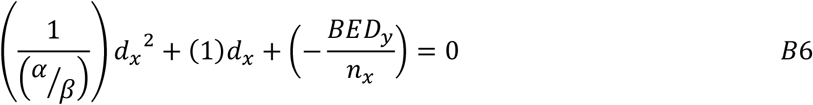

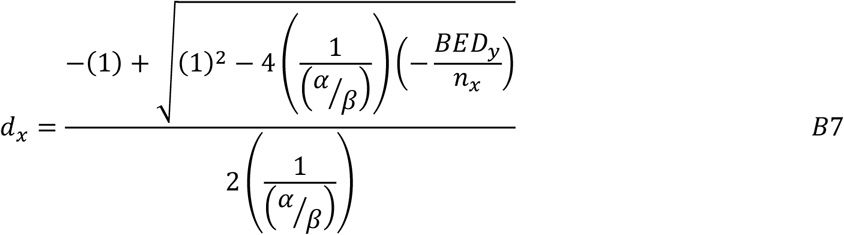

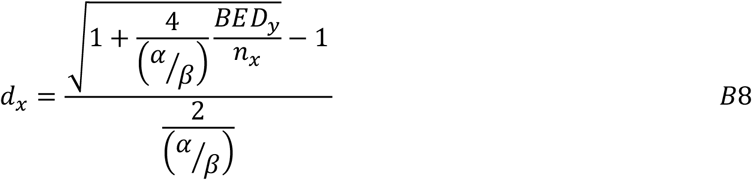

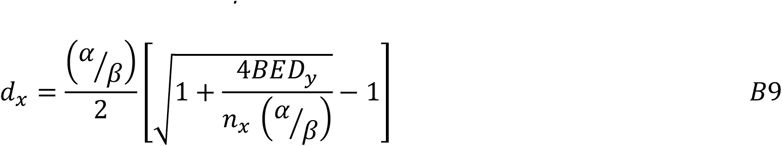

The total dose, i.e. equivalent physical dose (EPD), is then *D*_*x*_ = *n*_*x*_*d*_*x*_, which is given by

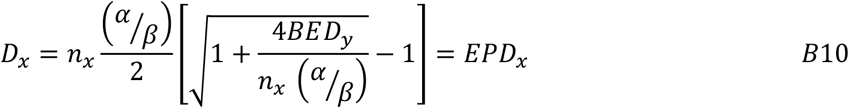

### Validation of equivalent biological effect

EPD can be directly substituted into the BED expression and results in an expression which can be reduced to identity, meaning that it holds true for all values of the variables involved.

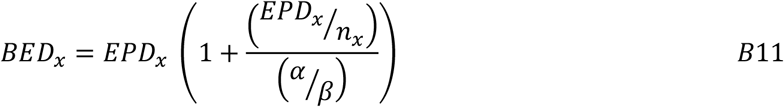

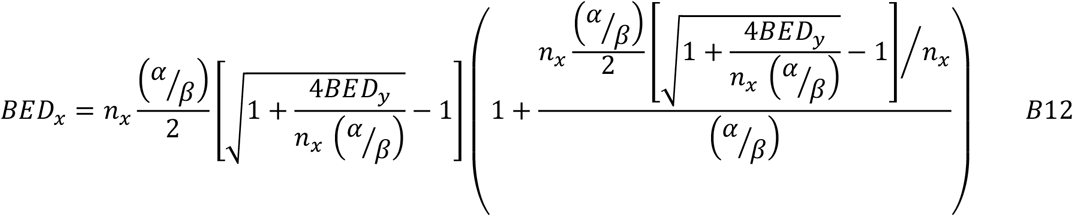

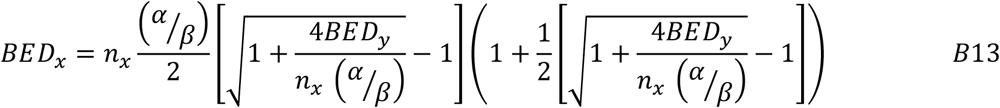

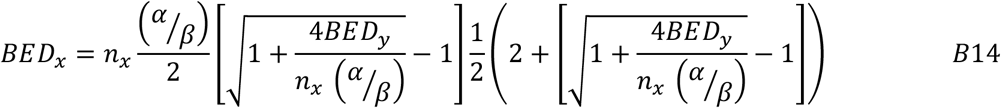

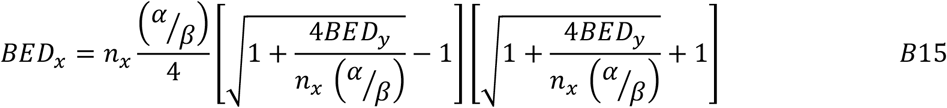

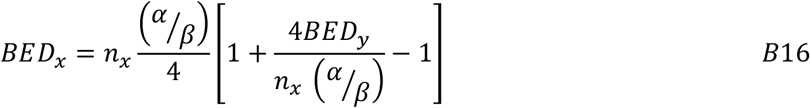

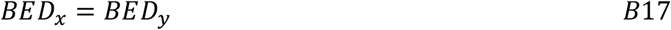

The inverse (or complement) of the BED operation, which moves BED back into physical dose space, correcting for biological effects according to the BED models, is demonstrated here. The units are the same for *EPD* and *D* but different for *BED*.

### Appendix C: Summary of Voxelized Formulae

This section summarizes the formulae used throughout the paper with variables that are consistently defined in simplest terms. Voxelized *BED* is straightforward to derive as it is calculated by linear-quadratic model dose scaling at each independent dose voxel *i* using

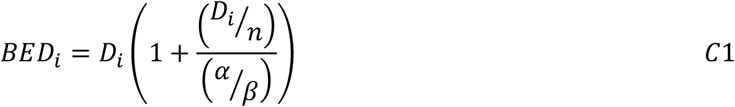

Equivalent dose in *x* Gy per fraction can be calculated for each voxel *i*. The voxelized *EQDx* equation implicit from the literature^26^ is

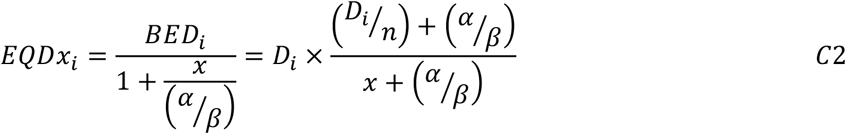

where *D*_*i*_ is the absorbed dose at voxel *i*. The *revised EQD* equation implicit from Proposition 1 is

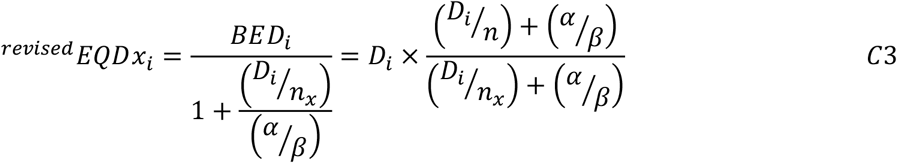

where *n*_*x*_ is the number of fractions of the reference therapy. The generalizeable voxelized equation per Proposition 2 has the generalized equivalent physical dose equation

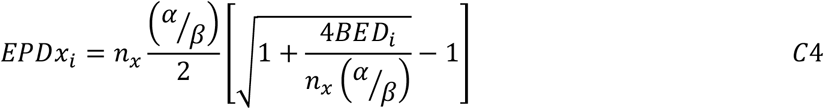

where *x* is the target prescription dose per fraction for the reference therapy, and *n*_*x*_ is the number of fractions of the reference therapy. In each of this preceding equations, equivalent dose in 2 Gy per fraction is obtained by substituting “2” in for the *x* of the voxelized equations.

## Acknowledgements

Acknowledgements

The authors thank Dr. Edward L. Sanders at Weatherall Institute of Molecular Medicine, Oxford University for his independent mathematics review and validation.

## Footnotes

* Note, *D*/*n* is intentionally used here for dose per fraction instead of *d* to emphasize that the total dose information *D*, unlike the quoted reference dose per fraction, is used directly in the voxelized dose distribution when performing treatment planning.

† Note, *D*/*n* is intentionally used here for dose per fraction instead of *d* to emphasize that the total dose information *D*, unlike the quoted reference dose per fraction *d*, is directly used in the voxelized dose distribution when performing treatment planning.

